# Metagenomic identification of severe pneumonia pathogens with rapid Nanopore sequencing in mechanically-ventilated patients

**DOI:** 10.1101/19002774

**Authors:** Libing Yang, Ghady Haidar, Haris Zia, Rachel Nettles, Shulin Qin, Xiaohong Wang, Faraaz Shah, Sarah F. Rapport, Themoula Charalampous, Barbara Methé, Adam Fitch, MS Alison Morris, Bryan J. McVerry, Justin O’Grady, Georgios D. Kitsios

## Abstract

**Background:** Metagenomic sequencing of respiratory microbial communities for etiologic pathogen identification in pneumonia may help overcome the limitations of current culture-based methods. We examined the feasibility and clinical validity of rapid-turnaround metagenomics with Nanopore™ sequencing of respiratory samples for severe pneumonia diagnosis.

**Methods and Findings:** We conducted a case-control study of mechanically-ventilated patients with pneumonia (nine culture-positive and five culture-negative) and without pneumonia (eight controls). We collected endotracheal aspirate samples (ETAs) and applied a microbial DNA enrichment method prior to performing metagenomic sequencing with the Oxford Nanopore MinION device. We compared Nanopore results against clinical microbiologic cultures and bacterial 16S rRNA gene sequencing. In nine culture-positive cases, Nanopore revealed communities with low alpha diversity and high abundance of the bacterial (n=8) or fungal (n=1) species isolated by clinical cultures. In four culture-positive cases with resistant organisms, Nanopore detected antibiotic resistance genes corresponding to the phenotypic resistance identified by clinical antibiograms. In culture-negative pneumonia, Nanopore revealed probable bacterial pathogens in 1/5 cases and airway colonization by *Candida* species in 3/5 cases. In controls, Nanopore showed high abundance of oral bacteria in 5/8 subjects, and identified colonizing respiratory pathogens in the three other subjects. Nanopore and 16S sequencing showed excellent concordance for the most abundant bacterial taxa.

**Conclusion:** We demonstrated technical feasibility and proof-of-concept clinical validity of Nanopore metagenomics for severe pneumonia diagnosis, with striking concordance with positive microbiologic cultures and clinically actionable information offered from the sequencing profiles of culture-negative samples. Prospective studies with real-time metagenomics are warranted to examine the impact on antimicrobial decision-making and clinical outcomes.

## Introduction

Pneumonia is a primary cause of morbidity and mortality among adults, leading to more than one million hospitalizations every year and high rates of intensive care unit (ICU) admission in the US (1). The mainstay of pneumonia management is early and appropriate antimicrobial therapy targeting the causative pathogens, balanced with preventing antibiotic overuse and emergence of resistance (2). Thus, timely and accurate identification of causal pathogens is imperative yet remains challenging due to reliance on culture-based methods with low sensitivity and long turnaround times (48∼72 hours to actionable results (3)). Recently developed rapid polymerase-chain reaction (PCR) tests represent a significant advancement in the field (4), but these tests can only detect the presence/absence of selected panels of pathogens, and thus are not comprehensive enough in breadth or resolution. Culture-independent methods using next-generation sequencing of microbial communities may help overcome the limitations of current diagnostic testing (5–7).

Our group and others have provided proof-of-concept evidence that sequencing of the bacterial 16S rRNA gene (16S sequencing) in clinical respiratory specimens can provide diagnostic insights beyond standard microbiologic cultures (5,8–10). Nevertheless, standard 16S sequencing is not clinically applicable due to limited resolution (providing only genus-level bacterial identification) and lengthy sample processing, library preparation and data acquisition timelines (11). The advent of Nanopore metagenomic sequencing (Oxford Nanopore Technologies [ONT], UK) has offered unprecedented capacities for real-time, detailed profiling of microbial communities at species level (including viruses and fungi) (12–15). With recent technical improvements to overcome the high amounts of contaminating human DNA in clinical respiratory samples (16), Nanopore metagenomics may allow for the development of a novel diagnostic approach for pneumonia.

In this study, we sought to evaluate the technical feasibility and clinical validity of Nanopore metagenomic sequencing for etiologic diagnosis of severe pneumonia in mechanically-ventilated patients in the ICU.

## Materials and Methods

Detailed methods are provided in the Supplement.

### Study design and Participants

From June 2018 – March 2019, we carried out a nested case-control study from an ongoing registry enrolling mechanically-ventilated adult patients with acute respiratory failure in the Medical Intensive Care Unit (MICU) at the University of Pittsburgh Medical Center (UPMC) (5,17). Exclusion criteria included inability to obtain informed consent, presence of tracheostomy, or mechanical ventilation for more than 72 hours prior to enrollment.

We diagnosed clinical pneumonia based on consensus committee review of clinical, radiographic, and microbiologic data per established criteria (18). We selected 14 subjects with a clinical diagnosis of pneumonia (nine with culture-confirmed diagnosis [culture-positive pneumonia group] and five with negative cultures [culture-negative pneumonia group]). Culture-positivity was deemed when a probable respiratory pathogen was isolated in clinical microbiologic cultures of respiratory specimens obtained at the discretion of treating physicians (sputum, endotracheal aspirate [ETA], or bronchoalveolar lavage fluid [BALF]). In culture-positive cases, antibiotic susceptibility testing was done as per standard practice at UPMC’s clinical microbiology laboratory, and results were interpreted based on Clinical and Laboratory Standards Institute criteria (19). We also included eight subjects (controls) who did not have evidence of lower respiratory tract infection and were intubated either for airway protection (n=5) or respiratory failure from cardiogenic pulmonary edema (n=3). From enrolled subjects, we collected ETAs for research purposes (sequencing) within the first 48hr from intubation. In two subjects, we utilized ETAs obtained on the fifth day post-intubation (instead of their baseline samples) when there was clinical suspicion of ventilator-associated pneumonia (VAP) (Case 10 and 15). From plasma samples taken at the same time with ETAs, we measured plasma procalcitonin levels (17). We recorded demographic, physiological, and laboratory variables at the time of sample acquisition, from which we calculated clinical pulmonary infection scores (CPIS) (20), and reviewed the antimicrobial therapies administered for the first 10 days from intubation.

This study was approved by the University of Pittsburgh Institutional Review Board (Protocol PRO10110387). Written informed consent was provided by all participants or their surrogates in accordance with the Declaration of Helsinki.

### Microbial DNA sequencing approaches

We focused our sequencing approach on DNA-based organisms (i.e. excluding RNA viruses) and aimed to perform agnostic profiling for microbes (bacteria and fungi) present in the ETAs obtained from the patients in the ICU. However, metagenomic microbial DNA sequencing in clinical respiratory samples is technically challenging because of the high amounts of contaminating human DNA that can overwhelm the sequencing output (ratio of human:microbial DNA >99:1 (21). Therefore, we applied a human DNA depletion step in ETA samples that utilized a detergent-based (saponin) method for selective lysis of human cells and digestion of human DNA with nuclease, as recently described (16). We extracted genomic DNA with the DNeasy Powersoil Kit (Qiagen, Germantown, MD) and assessed the efficiency of human DNA depletion by comparing quantitative PCR (qPCR) cycle threshold (Ct) of a human gene (Glyceraldehyde 3-phosphate dehydrogenase - GAPDH) and the bacterial 16S rRNA gene (V3-V4 region) (22) between samples subjected to depletion vs. not (depleted vs. undepleted samples).

From extracted DNA in depleted samples, we prepared metagenomic sequencing libraries with a Rapid PCR Barcoding Kit (SQK-RPB004) and then ran on the MinION device (Oxford Nanopore Technologies (ONT), UK, UK) for five hours. We basecalled the output (i.e. converted the sequencing device output into nucleic acid base sequences) with the Guppy software and used the ONT platform, EPI2ME, for quality control, species identification [What’s In My Pot (WIMP) pipeline] and antimicrobial resistance gene analyses [ARMA workflow]. Samples that generated fewer than 300 high-quality microbial reads were excluded from further analyses. As an internal quality control for the reliability and reproducibility of Nanopore sequencing, we performed sequencing on two samples with extracted DNA from a mock microbial community with known composition (ZymoBIOMICS Microbial Community Standard) and compared derived vs. expected abundance of microbial species.

To further validate the results of Nanopore sequencing for bacterial DNA, we performed standard 16S rRNA gene (V4 region) PCR amplification and sequencing on the Illumina MiSeq Platform as a reference method for bacterial DNA sequencing (23). We processed 16S sequences using an in-house pipeline developed by the University of Pittsburgh Center for Medicine and the Microbiome (CMM) (24–29). Samples that generated fewer than 100 bacterial reads were excluded from further analyses.

### Ecological and statistical analyses

From sequencing reads obtained from Nanopore and 16S sequencing, we calculated alpha diversity by Shannon index, performed permutational multivariate analysis of variance (PERMANOVA) testing to assess compositional differences between sample types, and visualized compositional dissimilarities between samples with the non-metric multidimensional scaling (NMDS) method using the Bray-Curtis index. All analyses were performed with the R *vegan* package (30).

## Results

### Cohort description

We enrolled 22 mechanical-ventilated patients with acute respiratory failure: nine with retrospective consensus diagnosis of culture-positive pneumonia, five with culture-negative pneumonia, and eight controls. Clinical characteristics and outcomes for the three groups are shown in Table 1. Cases with culture-positive pneumonia had significantly higher CPIS and a trend for higher procalcitonin levels compared to controls (Table 1, Fig S1). At the time of enrollment, empiric antibiotics had been prescribed for all 14 patients with clinical diagnosis of pneumonia, as well as for 5/8 of control patients (Table 1, Fig S3).

**Table 1:** Characteristics of enrolled patients. Continuous variables are presented as medians (with interquartile ranges), and categorical variables are presented as N (%).

|  | Culture-Positive<br>Pneumonia | Culture-Negative<br>Pneumonia | Controls |
| --- | --- | --- | --- |
| N | 9 | 5 | 8 |
| Age, median [IQR], yrs | 58.3 [55.2, 62.6] | 55.8 [45.7, 62.8] | 61.2 [51.9, 67.4] |
| Male, N (%) | 5 (55.6) | 2 (40.0) | 6 (75.0) |
| BMI, median [IQR] | 24.0 [21.5, 34.6] | 31.2 [25.6, 32.9] | 28.1 [25.1, 36.1] |
| SOFA Score, median [IQR]* | 6.0 [4.0, 6.0] | 7.0 [6.0, 7.0] | 5.0 [4.0, 8.0] |
| PaO2:FIO2 ratio, median [IQR], mmHg | 158.0 [137.0, 275.0] | 150.0 [121.0, 208.0] | 221.5 [205.0, 319.5] |
| Heart rate (median [IQR]), beats per minute | 107.0 [92.0, 117.0] | 83.0 [82.0, 88.0] | 81.5 [73.8, 85.5] |
| SBP (median [IQR]) | 125.0 [102.0, 141.0] | 118.0 [117.0, 127.0] | 105.0 [97.8, 117.5] |
| WBC, median [IQR], x 10<sup>9</sup> per liter (L) | 10.0 [7.4, 16.8] | 4.6 [3.5, 8.3] | 5.4 [4.5, 11.6] |
| Platelets, median [IQR], x 10<sup>9</sup> per liter (L) | 190.0 [169.0, 281.0] | 155.0 [136.0, 210.0] | 141.0 [72.2, 171.0] |
| Creatinine, median [IQR], mg/dL | 1.3 [1.1, 1.5] | 1.5 [1.5, 2.0] | 1.0 [0.8, 1.4] |
| Respiratory Rate, median [IQR], 1/min | 22.0 [21.0, 24.0] | 21.0 [20.0, 24.0] | 17.0 [15.5, 17.2] |
| PEEP, median [IQR], cm | 8.0 [5.0, 8.0] | 5.0 [5.0, 8.0] | 5.0 [5.0, 5.8] |
| Tidal Volume (per kg of PBW), (median [IQR]), ml/kg | 6.8 [6.2, 8.4] | 6.2 [6.1, 6.6] | 6.3 [6.0, 7.1] |
| Plateau Pressure, median [IQR], cm | 20.0 [13.0, 23.5] | 25.0 [21.0, 29.0] | 16.0 [13.0, 21.5] |
| Ventilator free days, median [IQR], days | 12.0 [0.0, 23.0] | 17.0 [6.0, 23.0] | 24.5 [24.0, 26.0] |
| ICU Length of Stay, median [IQR], days | 8.0 [5.0, 18.0] | 11.0 [6.0, 12.0] | 4.5 [3.8, 5.0] |
| Acute Kidney Injury, N (%) | 8 (88.9) | 4 (80.0) | 2 (25.0) |
| 30 Day Mortality, N (%) | 3 (33.3) | 1 (20.0) | 1 (12.5) |
| On antibiotic therapy, N (%) | 9 (100) | 5 (100) | 5 (62.5) |
| Antibiotic days, median [IQR], days | 12.0 [10.0, 24.0] | 21.0 [20.0, 24.0] | 3.5 [0.0, 7.5] |
| CPIS, median [IQR], | 8.0 [7.0, 9.0] | 6.0 [5.0, 7.0] | 5.0 [4.0, 5.2] |
| Procalcitonin, median [IQR], pg/μl | 2783.0 [1049.5, 4330.1] | 4866.0 [94.4, 4965.1] | 353.5 [250.4, 1531.6] |

Abbreviations: IQR: interquartile range; BMI: body mass index; SOFA: sequential organ failure assessment; PaO<sub>2</sub>: partial pressure of arterial oxygen; FiO<sub>2</sub>: fractional inhaled concentration of oxygen; SBP: systolic blood pressure; WBC: white blood cell count; PEEP: positive end-expiratory pressure; PBW: predicted body weight; ICU: intensive care unit; CPIS: clinical pulmonary infection score.
\* SOFA score calculation does not include the neurologic component of SOFA score because all patients were intubated and receiving sedative medications, impairing our ability to perform assessment of the Glasgow Coma Scale in a consistent and reproducible fashion.

### Technical feasibility of Nanopore sequencing in clinical samples

Pre-processing of the ETA samples with the saponin-based human DNA depletion protocol resulted in relative enrichment of bacterial DNA by an average of 1260-fold (Fig S2). This microbial enrichment step allowed for generation of sufficient numbers of microbial reads by Nanopore sequencing in depleted samples (median 6682 reads, average proportion 48% of total reads), whereas in undepleted samples the sequencing output was overwhelmed by human DNA (only 1% of reads were of microbial origin) and effectively was unusable (Fig 1A). Importantly, the depletion protocol did not appear to alter the underlying bacterial communities, because ecological analyses of depleted and undepleted samples by 16S rRNA gene sequencing demonstrated no significant differences in alpha or beta diversity (Fig 1B, C).

**Fig 1.**
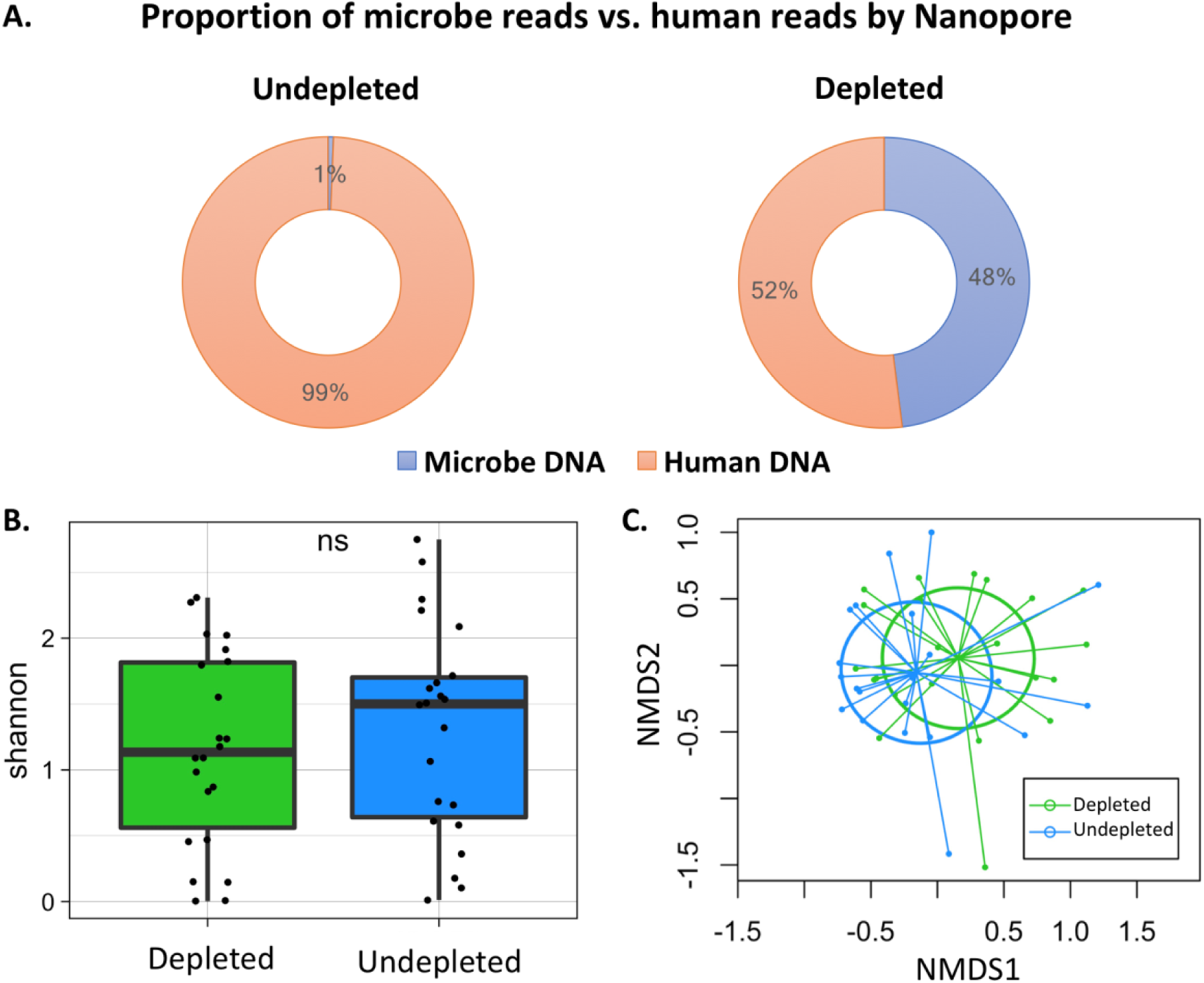
Saponin-based human DNA depletion effectively removed human DNA without changing bacterial community structure. (A) Before human DNA depletion, 1% of Nanopore reads were of microbial origin; following human DNA depletion, 48% of Nanopore reads were of microbial origin microbes. (B) There was no significant difference in alpha diversity of bacterial communities between depleted and undepleted samples assessed by 16S rRNA gene sequencing. (C) Non-metric multidimensional scaling (NMDS) plot of the Bray-Curtis dissimilarity index between depleted and undepleted samples based on 16S rRNA gene sequencing. Depleted samples were compositionally similar to undepleted samples (PERMANOVA, p-value=0.17).

### Analytical validity of Nanopore sequencing

Nanopore-derived bacterial communities showed striking similarity with both mock communities of extracted DNA (table S1) as well as 16S-derived community profiles for bacteria from clinical samples (Fig S3), underscoring the analytical validity of Nanopore results for use in further analyses.

### Nanopore community profiles by clinical group

By Nanopore sequencing, culture-positive samples had a trend for lower alpha diversity (Shannon index) compared to culture-negative samples (Fig 2A and Fig S3A) and demonstrated global compositional dissimilarities compared to culture-negative and control samples (PERMANOVA p-value=0.038, R^2^=0.12, Fig 2B and Fig S3B).

**Fig 2.**
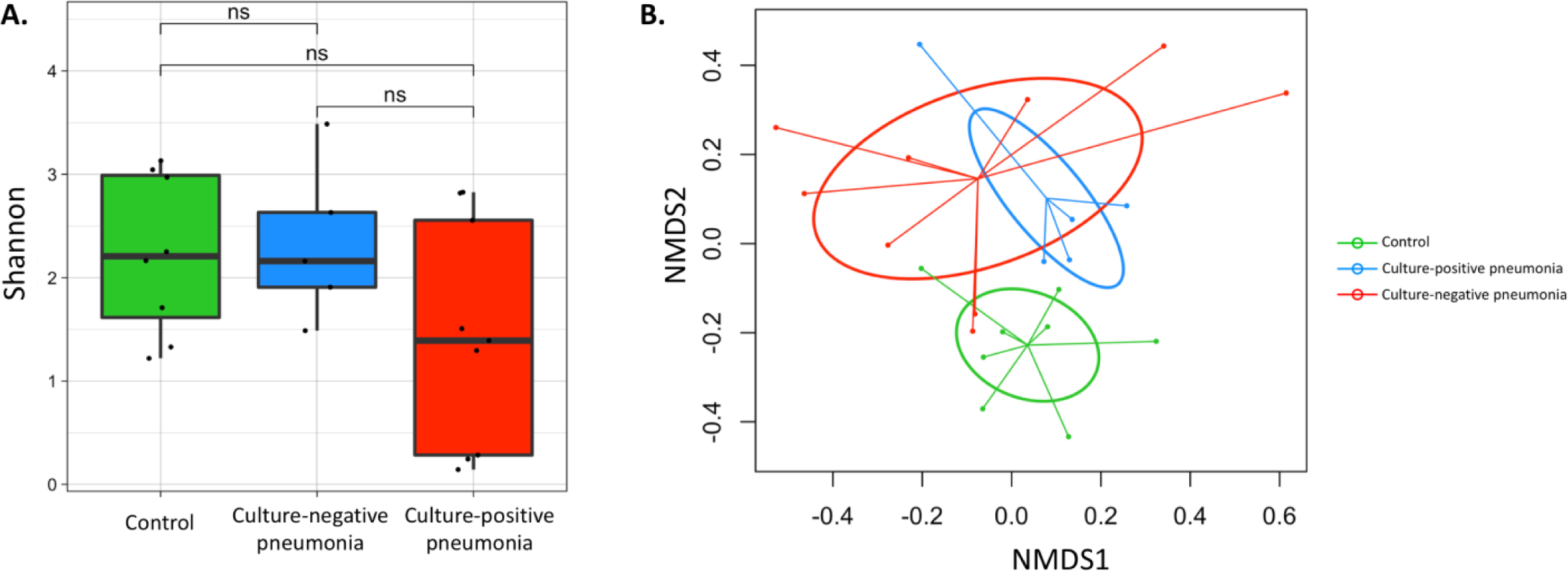
Comparisons of lung microbial communities between samples of culture-positive pneumonia, culture-negative pneumonia and controls based on Nanopore metagenomic sequencing. (A) Compared to samples from patients with culture-negative pneumonia, culture-positive samples had a trend for lower alpha diversity of lung microbial communities by Shannon index. (B) By non-metric multidimensional scaling (NMDS) plot of the Bray-Curtis dissimilarity index, there were significant differences in overall microbial community compositions between three groups (PERMANOVA for Bray-Curtis dissimilarity index, p-0.0378, R^2^=0.120).

### Nanopore-based pathogen identification

#### Culture-positive pneumonia

We first examined Nanopore results in the culture-positive cases in which microbiologic confirmation of the causative pathogen allowed for a targeted interrogation of the sequencing output for the corresponding microbial species. We examined different thresholds of sequencing output (i.e. absolute number of reads for the dominant pathogen vs. relative or ranked abundance thresholds for pathogens) to maximize sensitivity of Nanopore results for detecting the culture-identified pathogen(s). By focusing on the three most abundant species (bacterial or fungal) by Nanopore sequencing, we were able to identify all culture-confirmed pathogens with high relative abundances.

In eight culture-positive bacterial pneumonias, Nanopore profiles showed high abundance of the same bacterial species isolated in cultures (Fig 3A). These highly abundant causative bacterial pathogens had on average 90 times higher relative abundance compared to the species ranked second in abundance in each community (Fig 3B). Nanopore sequencing also revealed high abundance of additional potential bacterial pathogens in 2/8 of samples that were not detected by cultures (*E. coli* in subject 1 and *H. influenzae* in subject 8), suggesting the presence of a polymicrobial infection despite the isolation of a single pathogenic bacterial species on standard cultures (Fig 3A).

**Fig 3.**
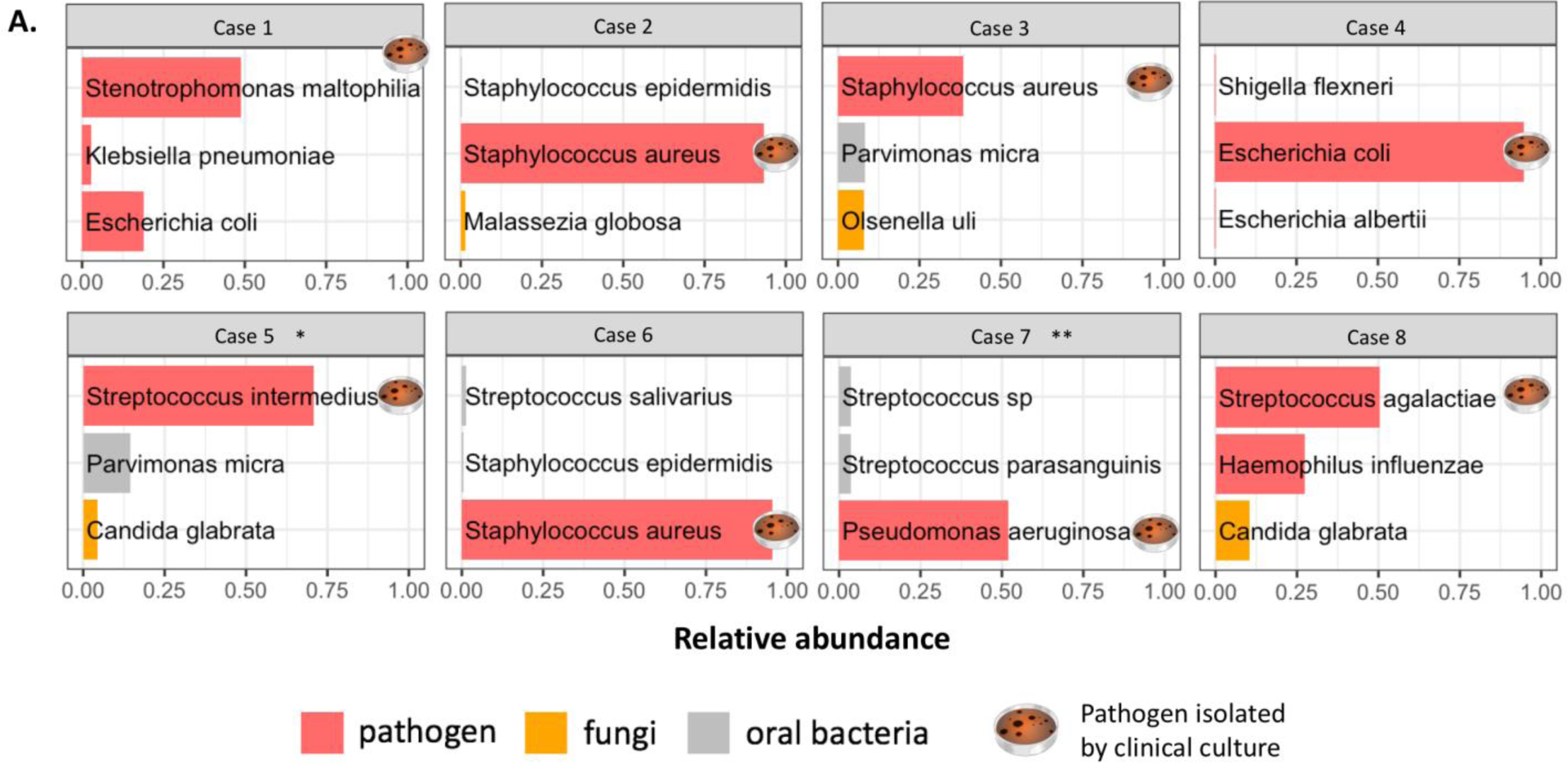
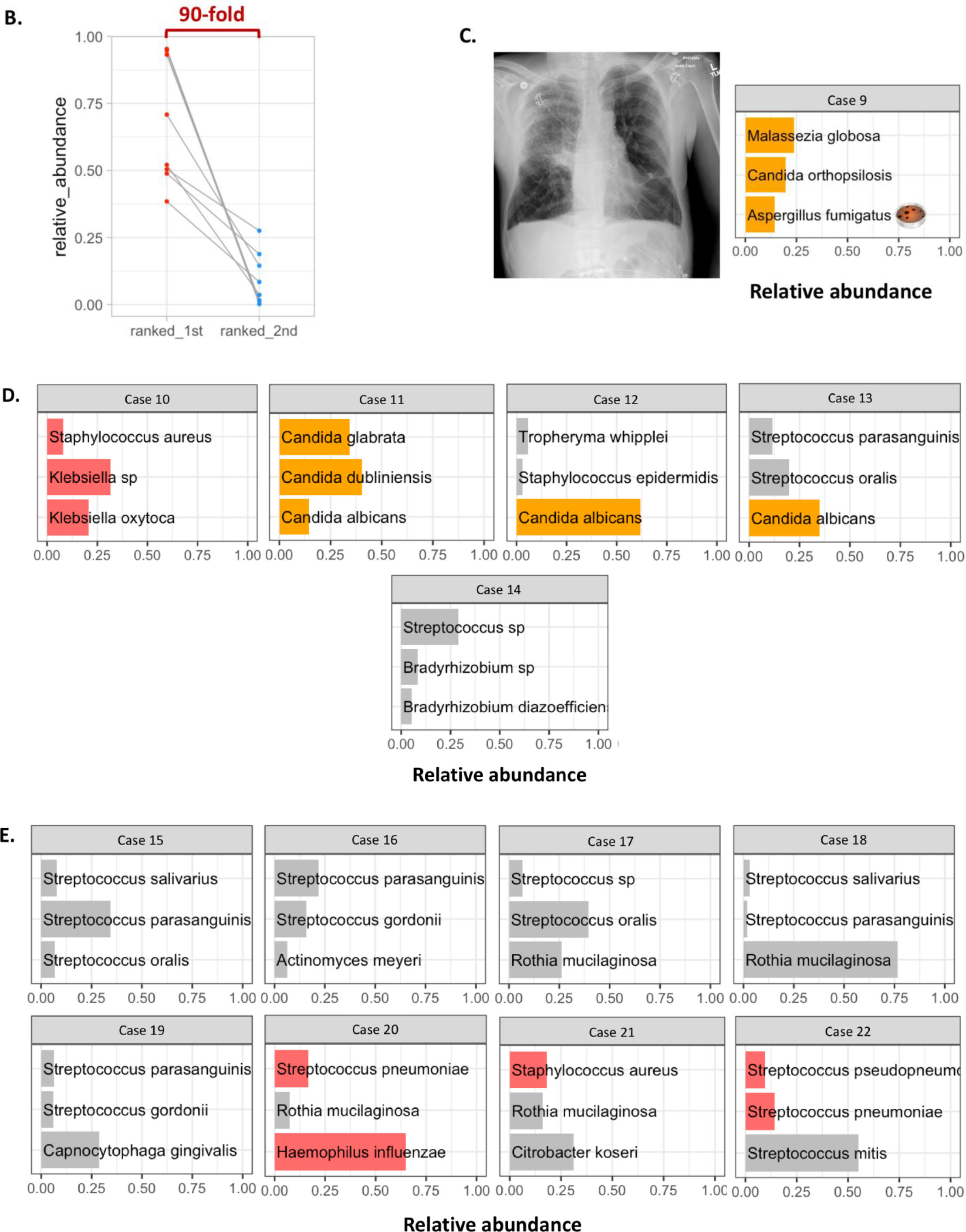
Comparisons of microbes detected by Nanopore metagenomic sequencing and clinical culture. Each small plot represents an endotracheal aspirate; Each bar represents a microbe; the X-axis represents the relative abundance of microbes by Nanopore. Petri dish represents pathogen isolated by clinical culture. The three most abundant taxa detected by Nanopore sequencing were included. (A) In 8 samples with culture-positive bacterial pneumonia, Nanopore signals were dominated by pathogens isolated by culture. (B) In 8 samples with culture-positive bacterial pneumonia, the relative abundance of culture-positive pathogens was 90-times higher than that of the second-ranked taxa detected by Nanopore. (C) In 1 sample with probable invasive fungal infection, chest radiograph supported a clinical diagnosis of pneumonia, *Aspergillus fumigatus* was isolated by culture, and Nanopore revealed the same fungal pathogen by sequencing. (D) In 5 culture-negative pneumonia samples, potential pathogens were found in one sample, and fungi were found in 3 samples with Nanopore. (E) Only typical oral bacteria were identified in 5/8 of control samples, but potential pathogens were detected in 3/8 of them. * compared to culture of pleural fluid; ** case of culture-positive tracheobronchitis and acute exacerbation of chronic obstructive pulmonary disease (no infiltrate on chest radiograph)

These eight culture-positive cases with clinical antibiograms allowed for examination of the potential predictive utility of antibiotic resistance gene detection with metagenomic sequencing (Table 2). In the single case of methicillin-resistant *Staphylococcus aureus* (MRSA, case 2, Fig S3), Nanopore detected 4 reads aligned to the responsible *mecA* gene, whereas in the three cases of methicillin-sensitive *S. aureus* (MSSA, cases 3, 5 and 6), no *mecA* gene reads were detected. Similarly, in the two cases of *Stenotrophomonas maltophilia* and *E*.*coli*, Nanopore detected genes that explained the observed phenotypic antimicrobial resistance profile (Table 2).

**Table 2:** Comparison of antibiotic resistance phenotype detected by clinical culture and clinically relevant resistance genes detected by Nanopore in cases of bacterial pneumonia. Genes conferring resistance phenotype are highlighted in bold.

| Sample_ID | Pathogen by culture & Nanopore | Resistance phenotype by culture | Clinically relevant resistance genes [N of alignments] |
| --- | --- | --- | --- |
| Resistance identified |  |  |  |
| Case 1 | *Stenotrophomonas maltophilia* | R: ticarcillin/clavulanic acid<br>R: ceftazidime | *blaTEM-4* [4]<br>*blaTEM-112* [1]<br>*blaTEM-157* [1]<br>*blaACT-5* [1] |
|  |  | I: levofloxacin | *oqxB* [1] |
|  |  | Tetracycline not tested | *tetC* [6] |
| Case 2 | *Staphylococcus aureus* | R: methicillin | *mecA* [4] |
|  |  | R: erythromycin, clindamycin | *ermA* [10]<br>*erm(33)* [1] |
|  |  |  | *tet38* [11]<br>*ant(4')-Ib* [9]<br>*tetC* [3]<br>*blaTEM-4* [1] |
| Case 3 | *Staphylococcus aureus* | I: tetracycline | *tetK* [1]<br>*tet38* [1]<br>*tetQ* [1] |
| Case 4 | *Escherichia Coli* | R: tetracycline | *tetX* [1] |
|  |  | R: Trimethoprim-sulfamethoxazole | *sul1* [363]<br>*dfrA* [127] |
|  |  | R: ciprofloxacin, levofloxacin | *acrF* [315]<sup>§</sup><br>*parE* [304]<sup>§</sup><br>*mfd* [277]<sup>§</sup> |
|  |  |  | *mphA* [215]<br>*aadA5* [196]<br>*vgaC* [110]<br>*blaACT-5* [7]<br>*blaACT-14* [3]<br>*mefA* [1]<br>*mel* [1] |
| No resistance identified |  |  |  |
| Case 5 | *Staphylococcus aureus* | S: all tested agents | none |
| Case 6 | *Staphylococcus aureus* | S: all tested agents | *tet38* [2]<br>*blaTEM4* [2] |
| Case 7 | *Pseudomonas aeruginosa* | S: all tested agents | none |
| Resistance not tested |  |  |  |
| Case 8 | *Streptococcus agalactiae* | Not tested | *tetM* [60]<br>*isaC* [55]<br>*sul1* [2]<br>*tetQ* [2]<br>*mphA* [1]<br>*aadA5* [1] |

Abbreviations: R: resistant; I: intermediate; S: Susceptible
§ Genes conferring antibiotic resistance phenotype but not classified as clinical relevant genes by EPI2ME antimicrobial resistance gene analyses [ARMA workflow]
Tested agents for case 5: Ampicillin/Sulbactam, Oxacillin, Imipenem, Gentamicin, Erythromycin, Tetracycline, Vancomycin, Clindamycin, Linezolid, Rifampin, Trimethoprim-sulfamethoxazole, Synercid.
Tested agents for case 6: Ampicillin/Sulbactam, Oxacillin, Imipenem, Gentamicin, Erythromycin, Tetracycline.
Tested agents for case 7: Piperacillin/Tazobactam, Ticarcillin/Clavulanic acid, Cefepime, Ceftazidime, Imipenem, Meropenem, Aztreonam, Gentamicin, Tobramycin, Amikacin, Ciprofloxacin, Levofloxacin.

We also tested the performance of Nanopore sequencing in one case with probable invasive fungal infection. Subject 9 was a lung transplant recipient who had been receiving antifungal therapy for a positive sputum culture for *Aspergillus fumigatus*. Clinical decompensation with acute respiratory failure raised concern for bacterial co-infection and initiation of intensive broad-spectrum antibiotics. BALF culture grew again *Aspergillus fumigatus*, which was the dominant pathogen detected by Nanopore, without any other sequencing evidence of bacterial infection (Fig 3C). Thus, in this case with probable invasive fungal infection, Nanopore sequencing only identified a confirmed fungal pathogen and ruled out the presence of bacterial pneumonia.

#### Culture-negative pneumonia

Nanopore sequencing provided diverse representations of microbial communities in cases of clinical suspicion of pneumonia with negative cultures, and thus interpretation needs to be individualized for each case (Fig S3).

In case 10 with an initial diagnosis of aspiration pneumonia caused by *S. aureus* and *Klebsiella oxytoca* identified by BALF culture on day 2 post-intubation, clinical deterioration on appropriate antibiotic therapy and new radiographic infiltrates by day 5 raised concern for VAP. However, repeat BAL culture on day 5 was negative. Nanopore detected high abundance of both *S. aureus* and *Klebsiella oxytoca* on day 5 sample, revealing that the culture-negative community consisted of abundant previously identified pathogens, which were likely not viable at the time of the day 5 BALF acquisition due to ongoing antibiotic therapy. Moreover, low procalcitonin level at the time of the day 5 sample (94 pg/μl) and absence of new pathogens by sequencing made diagnosis of new VAP unlikely.

In another case of a lung transplant recipient (subject 11) with diffuse bilateral consolidations and persistent clinical septic picture of undefined etiology, BALF culture was only positive for yeast and the patient was empirically treated with broad-spectrum antibiotics. Eventually, the patient was proven to be fungemic with delayed growth of *Candida glabrata* on initial blood cultures prompting addition of antifungal therapy. Nanopore sequencing on ETA sample from day 1 post-intubation showed high abundance of *Candida glabrata* and *Candida dubliniensis* with very low abundance of bacterial reads, confirming the absence of bacterial pneumonia and demonstrating fungal colonization of the allograft. Of note, in two other culture-negative cases, Nanopore also detected high abundance of *Candida albicans* (Fig 3D), whereas in the last case, both Nanopore and 16S sequencing identified abundant oral bacteria with no typical respiratory pathogens.

#### Controls

Eight control subjects did not meet clinical diagnostic criteria for pneumonia on retrospective examination of their clinical course. Despite not meeting diagnostic criteria, 5/8 cases empiric antibiotics were prescribed empiric antibiotics early in their course for initial consideration of possible pneumonia. Five samples were dominated by common oral bacteria, such as *Rothia* and non-*pneumoniae Streptococcus* species (31,32). However, in the other three samples, Nanopore and 16S sequencing detected potential respiratory pathogens (e.g. *S*.*aureus, H. influenzae* or *S. pneumoniae*) that were likely airway colonizers not causing clinical infection, notion supported by clinical improvement despite early discontinuation of antibiotics and/or low procalcitonin levels (Fig S3). No significant fungal DNA presence was detected by Nanopore sequencing in the control group (Fig 3E).

## Discussion

In this nested case-control study, we provide proof-of-concept evidence that untargeted, shotgun metagenomic sequencing with the MinION device can provide clinically useful information for etiologic diagnosis of pneumonia in mechanically-ventilated patients. We demonstrate feasibility of metagenomic sequencing directly from clinical respiratory specimens by applying a saponin-based protocol for human DNA depletion prior to sequencing. Our analyses demonstrated global microbial community structure and species-level compositional di?erences associated with culture-positivity and clinical diagnosis of pneumonia. Nanopore sequencing had striking concordance with cultures by detecting high abundance of the causative pathogenic bacteria in culture-positive cases and refuting bacterial pneumonia diagnosis in selected culture-negative cases.

Nanopore metagenomic sequencing holds promise as a potential infection diagnostic tool due to its comprehensive scope, high resolution and real-time data generation (6). However, contaminating human DNA has been a rate-limiting step for clinical implementation of direct-from-sample sequencing in respiratory specimens. By applying a recently validated protocol with saponin-based, human DNA depletion (13), we demonstrate that this approach is feasible, reproducible and effective for maximizing the microbial signal in clinical samples and providing reaching reliable diagnostic output. With further technical improvements, process automation and cost reductions of metagenomic sequencing, this approach could be introduced in the clinical microbiology laboratories as part of the diagnostic pipeline.

Nanopore sequencing showed high accuracy in pathogen identification in culture-positive pneumonia. Obviating the need for *ex-vivo* growth for organisms, direct-from-sample sequencing can offer comprehensive snapshots of the component microbes of the communities at the time of sample acquisition. Sequencing methods are thus robust to specific growth condition requirements or the impaired viability of organisms due to antecedent antimicrobials, factors that can lead to the ‘great plate count anomaly’ of culture-based methods, i.e. the absent or limited growth of bacteria in cultures despite abundant visualized organisms on specimen gram-staining (33). In exploratory analyses of the sequencing output, Nanopore also provided antibiotic resistance information by detecting clinically relevant resistance genes that matched the phenotypic resistance on antibiograms (e.g. *mecA* detection/absence in MRSA/MSSA cases, respectively). Overall, rapid metagenomic sequencing closely matched the results of current, reference-standard diagnostic methods in our cohort, which typically take 2-3 days for allowing appropriate antibiotic adjustments to occur. Thus, with further external validation in additional cohorts, nanopore metagenomics hold promise for rapidly accelerating the diagnosis of etiologic pathogens and shortening the time to appropriate therapy selection in cases where diagnosis currently relies on cultures.

Invasive pulmonary fungal infections represent a major diagnostic challenge due to the poor sensitivity and slow turnaround times of cultures and the need for invasive samples with histopathology for diagnostic confirmation (34). In the single case of *Aspergillus fumigatus* infection, Nanopore confirmed the high abundance of *Aspergillus fumigatus* in the community and ruled out concomitant bacterial pneumonia. Such results can directly influence clinical practice, as unnecessary and potentially harmful antibiotics could be discontinued with antimicrobial therapies focused on the causative fungal pathogen (35).

In cases of culture-negative pneumonia and in controls, the main compositional pattern consisted of diverse communities with oral bacteria abundance (5), similar to clinical microbiology reports of normal respiratory flora. In such cases, early de-escalation or discontinuation of antibiotics could be further supported by sequencing results, if available in real-time. Nonetheless, in 3/5 airway controls, potential respiratory pathogens were detected in high abundance by both Nanopore and 16S sequencing, in the absence of other supportive evidence of pneumonia. Such cases highlight that the high sensitivity of sequencing for identifying pathogenic organisms missed by cultures could accentuate the existing clinical challenge of distinguishing colonizing vs. infecting organisms in the airways of mechanically-ventilated patients. In such scenarios, the distinction between colonization and infection cannot be based solely on microbial DNA sequencing outputs, but needs to be an integrative one, incorporating clinical, radiographic, and systemic or focal host-responses (36–38). At the same time, knowledge of colonizing organisms in critically-ill patients could facilitate more targeted choices for initial antibiotic regimens in the event of a secondary infection, such as VAP.

Our study is limited by the single center design and the available sample size. We did not perform Nanopore sequencing and data analyses in real-time because of our retrospective study design, and our objective of demonstrating proof-of-concept feasibility. Nonetheless, the method is implementable with short turnaround times (∼6-8hrs) (13). The results of antibiotic resistance gene sequencing should be interpreted with caution, given that our analyses were exploratory, included a limited number of multidrug-resistant bacteria that precluded a formal predictive modeling analysis. Thus, development of reliable predictive rules for pneumonia diagnosis or resistance gene identification based on sequencing outputs will require prospective examination of large cohorts of patient samples. Finally, the human DNA depletion method we applied is not yet optimized for viral DNA detection (16). Nonetheless, most clinically relevant respiratory viruses are RNA organisms, which are not within the scope of current DNA-based metagenomics, but can be detected with available PCR-based panels. With evolving rapid metagenomic platforms that can also sequence RNA molecules (39), the profiling of respiratory viruses as well as host transcriptomic responses (37) will enable more comprehensive representations of the altered respiratory ecosystem in pneumonia.

In conclusion, our study demonstrates the technical feasibility and clinical validity of direct-from-specimen metagenomics with a rapid protocol for human DNA depletion protocol and sequencing with the MiNION device. Metagenomic approaches hold promise for the development of rapid and comprehensive diagnostic tools for severe pneumonia that could transform the existing diagnostic paradigm. With real-time data generation and turnaround times of 6-8hrs from sample acquisition to result, rapid metagenomics could conceivably allow for targeted adjustment of initial empiric antibiotic regimens even before their second dose is due, and thus allow for personalized antimicrobial prescriptions and antibiotic stewardship gains. Our results provide strong rationale for a prospective, large-scale study with real-time application of metagenomics in order to measure the direct impact on antibiotic guidance and clinical outcomes.

Data Sharing Statements: All de-identified sequencing data were submitted to Sequence Read Archive (SRA) database, accession numbers 12268279 – 12268349. All de-identified datasets for this study are provided in https://github.com/MicrobiomeALIR.

## Data Availability

Data Sharing Statements: All de-identified sequencing data were submitted to Sequence Read Archive (SRA) database, accession numbers 12268279 - 12268349. All de-identified datasets for this study are provided in https://github.com/MicrobiomeALIR.

## Acknowledgement

We would like to thank all members of the research team of the Acute Lung Injury Registry (ALIR) and Biospecimen Repository at the University of Pittsburgh, the medical and nursing staff in the Medical Intensive Care Unit at the University of Pittsburgh Medical Center, and all patients and their families for participating in this research project.

## Notes

Conflicts of Interest: Dr. Bryan J. McVerry is a consultant for Vapotherm, Inc. Dr. Georgios Kitsios receives research funding from Karius, Inc. Dr. Justin O’Grady receives (or received) research funding and consumable support from Oxford Nanopore Technologies (ONT) and financial support for attending conferences and for speaking at ONT headquarters. The other authors have no conflicts of interest to declare.

### Competing Interest Statement

Dr. Bryan J. McVerry is a consultant for Vapotherm, Inc. Dr. Georgios Kitsios receives research funding from Karius, Inc. Dr. Justin O'Grady receives (or received) research funding and consumable support from Oxford Nanopore Technologies (ONT) and financial support for attending conferences and for speaking at ONT headquarters. The other authors have no conflicts of interest to declare.

### Funding Statement

Funding support: National Institutes of Health [K23 HL139987 (GDK); U01 HL098962 (AM); P01 HL114453 (BJM); R01 HL097376 (BJM); K24 HL123342 (AM); K23 GM122069 (FS)]. This paper presents independent research funded by the National Institute for Health Research (NIHR) under its Program Grants for Applied Research Program (reference no. RP-PG-0514-20018, JOG.), the UK Antimicrobial Resistance Cross Council Initiative (no. MR/N013956/1, JOG), Rosetrees Trust (no. A749, JOG) and the Biotechnology and Biological Sciences Research Council (BBSRC) Institute Strategic Programme Microbes in the Food Chain BB/R012504/1 and its constituent projects BBS/E/F/000PR10348 and BBS/E/F/000PR10349 (JOG).

### Author Declarations

All relevant ethical guidelines have been followed and any necessary IRB and/or ethics committee approvals have been obtained.

Any clinical trials involved have been registered with an ICMJE-approved registry such as ClinicalTrials.gov and the trial ID is included in the manuscript.

